# Prototyping a Generative AI-powered Person-centered Digital Health Tool to Mitigate Risk of Preventable Adverse Drug Events

**DOI:** 10.64898/2026.06.02.26354712

**Authors:** Duncan Dobbins, Angelique Russell, Megan Gunther, Vishal Shetty, Alexandra Shomali, David K. Vawdrey, Stephen C. Waring, Payton Whary, Jenna Wong, Eric A. Wright, Anthony W. Olson

## Abstract

**Objectives:** Older adults with comorbidities and polypharmacy have disproportionately high risk of hospitalization as well as readmission from adverse drug events (ADEs), of which 28%-71% are preventable (pADEs). This paper introduces an LLM application, CommunicADE, designed to support risk-mitigation of pADE-related readmission for the aforementioned population. We aim to evaluate CommunicADE’s technical performance with OpenAI’s HealthBench criteria: accuracy, completeness, communication quality, context awareness, and instruction following.

**Materials and Methods:** Our technical validation study used an LLM (KimiK2.5) to simulate interviews between CommunicADE and nine high-fidelity synthetic patients hospitalized and at increased risk for pADE-related readmission (65+ years, comorbidities, 5+ medications). Some pADE risk mechanisms clues were visible to CommunicADE in patient H&Ps, but most mechanisms were solely discoverable in interviews. Two pharmacists evaluated CommunicADE’s interview questions and EHR notes with HealthBench-informed variables. Analyzes used descriptive statistics.

**Results:** For 35 mechanisms across 9 patients (x□=3.89/patient), CommunicADE’s precision and recall were 0.92 and 0.63, respectively. Hallucinations were absent. Coherence and person-centeredness scored 4.28 and 4.44 on a 5-point scale (5=highest). On average, communication was at a 5^th^ grade level and objective for 78% of patients. Most patient-reported quotes included in notes (92%) supported detected mechanisms. CommunicADE followed all instructions regarding interview length and patient approvals.

**Discussion:** CommunicADE’s strongest performance was in accuracy (precision, hallucinations), communication quality (coherence, readability), context awareness (person-centeredness). Completeness (recall) and instruction following (objectivity, pADE mechanism/quote alignment) show room for improvement.

**Conclusion:** Findings suggest technical readiness for a feasibility pilot with real-world patients, and key areas for performance improvement.

## INTRODUCTION

### Background and Significance

Adverse drug events (ADEs) send 1.5 million Americans to emergency departments annually, of which one-third are hospitalized.^1^ ADEs are also the third leading cause of death in the United States behind cardiovascular diseases and cancer.^2^ ADEs cost the United States hundreds of billions of dollars annually from inpatient stays, readmissions, added treatments (including inappropriate prescription cascades), missed work days, and malpractice litigation.^3–6^ Among older adults with comorbidities, 28%-71% of ADEs are preventable (pADEs),^7–10^ defined as unintended harmful events from the normal medication use caused by preventable errors occurring between clinician prescribing and patient administration (e.g., misuse, nonadherence).^11^

Several modifiable determinants can mitigate pADE risk, especially those identifiable and addressable in patient-clinician interactions at the point of care (e.g., knowledge deficits; missing/outdated/inaccurate information; health literacy; polypharmacy; nonadherence/misuse).^12^ However, only 7%-14% of non-nosocomial pADEs and their underlying modifiable determinants are documented or detected in electronic health records (EHRs).^12^ Even if EHRs contain relevant pADE determinant information, connecting the dots is not straightforward (e.g., complex diagnostic information, dynamic medication histories/laboratory values, varying quality and completeness of notes) and clinicians can miss key contextual patient-reported information for mitigating pADE risk.^12,14–16^ Clinicians also face challenges like continually changing patient panels and numerous interruptions that hinder remembering, documenting, assessing causality, and making plans to address pADEs.^17^ Similarly, patients and their caregivers may be unclear about the information to share with clinicians about their medication use (i.e., when, how, what, and whether), especially in time-constrained appointments.^18–20^ Some patients may also be apprehensive about being judged, lectured, embarrassed, or a burden to clinicians by expressing medication-related concerns or asking questions.^19^

There is a need for tools and workflows that support clinicians to efficiently detect important and timely information at the point of care for addressing modifiable determinants of pADEs. Leveraging recent advances in artificial intelligence with large language models (LLMs),^21–22^ we designed a patient-centered digital health tool named CommunicADE to help address this need.

### CommunicADE Prototype Description

LLMs work by rapidly predicting the next word in a sequence of text using context from preceding words to generate coherent and contextually relevant text. Open-source LLM chatbots have been engineered for patient interaction to improve care quality, particularly in clinical communication and care coordination (e.g., decreased fear of judgment at intake/triage, increased feelings of safety at follow-up).^23–25^ Most patient-facing chatbots provide administrative assistance (e.g., scheduling) or deliver specialized services and expert information (e.g., education, health behavior promotion).^26–27^ We are unaware of any theory-informed chatbots designed to collect important contextual detail on the values, preferences, beliefs, goals, and patterns of medication use directly from patients, who are the experts on how prescriptions fit into their daily lives.^21,27,280^

CommunicADE is an application customized to support secondary prevention of pADEs for the patient population at highest risk for these events: older adults (≥65 years) with comorbidities and polypharmacy being treated with opioids, anticoagulants, or hypoglycemic-prone antidiabetic agents (i.e., insulins, sulfonylureas). CommunicADE can be deployed on devices like tablets and smart TVs during the hospital admission process (e.g., medication history workflows) for patients with a suspected ADE. The application conducts semi-structured, patient-centered medication interviews powered by an LLM that collects and identifies evidence-based mechanisms for reducing pADE risks - information often absent from or unclear within the EHR. CommunicADE then transcribes the interview and synthesizes a corresponding patient-friendly clinical note that is reviewable, revisable, and approved by the patient before transmission to the EHR for clinician review (**Figure 1**). CommunicADE intends to supplement clinician encounters, not replace them; thus, interviews and notes do not contain subjective inferences of clinical recommendations. Instead, CommunicADE aims to facilitate improvements in the timely collection, organization, and communication of important contextualized patient-reported medication use information to the clinician. Simultaneously, it may also help patients to feel heard, safe, and cared for, and enables shared decision making.^12,29–32^

**Figure 1.**
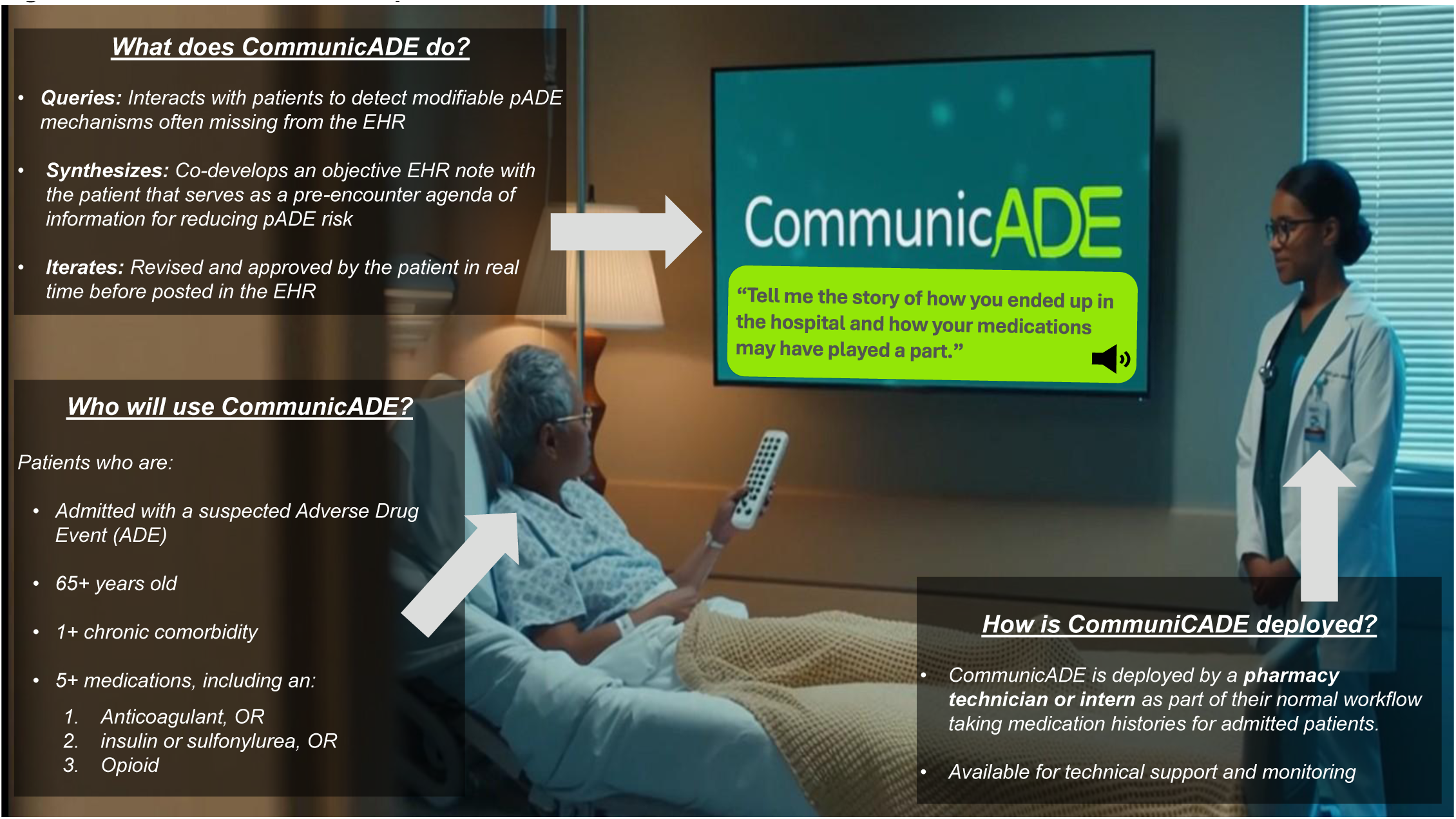
CommunicADE Description. **ALT TEXT:** An image containing descriptive text that showing what CommunicADE does as well as specifying its intended user and deployment in a pharmacy service workflow.

A robust three-step approach for validating the therapeutic benefit of LLM-powered healthcare chatbots like CommunicADE is: (1) technical validation; (2) pilot feasibility testing (confirm fidelity and acceptability); and (3) pragmatic effectiveness trials.^33^ This study’s objective is to validate CommunicADE’s technical performance with an Open-AI compatible LLM using five evaluation from OpenAI’s HealthBench (accuracy, completeness, quality of communication, context awareness, and instruction following).^34^

## METHODS

### Study Design and Data Source

Data for the technical performance validation were collected from text generated in interviews between CommunicADE and synthetic patients (i.e., digital representations of human patients generated using artificial intelligence to approximate the information, behaviors, and response of a real patient) using the LLM Kimi K2.5.^35^ LLMs have been previously used to conduct high-fidelity simulations clinical encounters with synthetic patients (i.e., realistic, medically accurate, appropriate empathy) preceding pilot feasibility and pragmatic trials in real-world clinical encounters.^36,37^ We followed seven applicable recommendations from the Conversational Reasoning Assessment Framework for Testing in Medicine (CRAFT-MD) to design the evaluation of CommunicADE, including: [R1] a dynamic conversation in a defined clinical setting (i.e., ADE-related hospitalization of a high-risk patient); [R2] use of open-ended questions; [R3] assessment of ability to gather essential information through conversation; [R5] bridging physician information gaps using LLMs (i.e., pADE risk mechanisms missing or hidden in EHR); [R7] refinement of prompting strategies to enhance prototype development; [R9] combining automated and expert evaluations; and [R10] inclusion of full prompt for transparency and transferability to development in other cases.^38^ **Figure 2** displays the multi-step process used with an OpenAI-compatible LLM to engineer CommunicADE, generate synthetic patients, simulate interviews, and the evaluate technical performance of CommunicADE.

**Figure 2.**
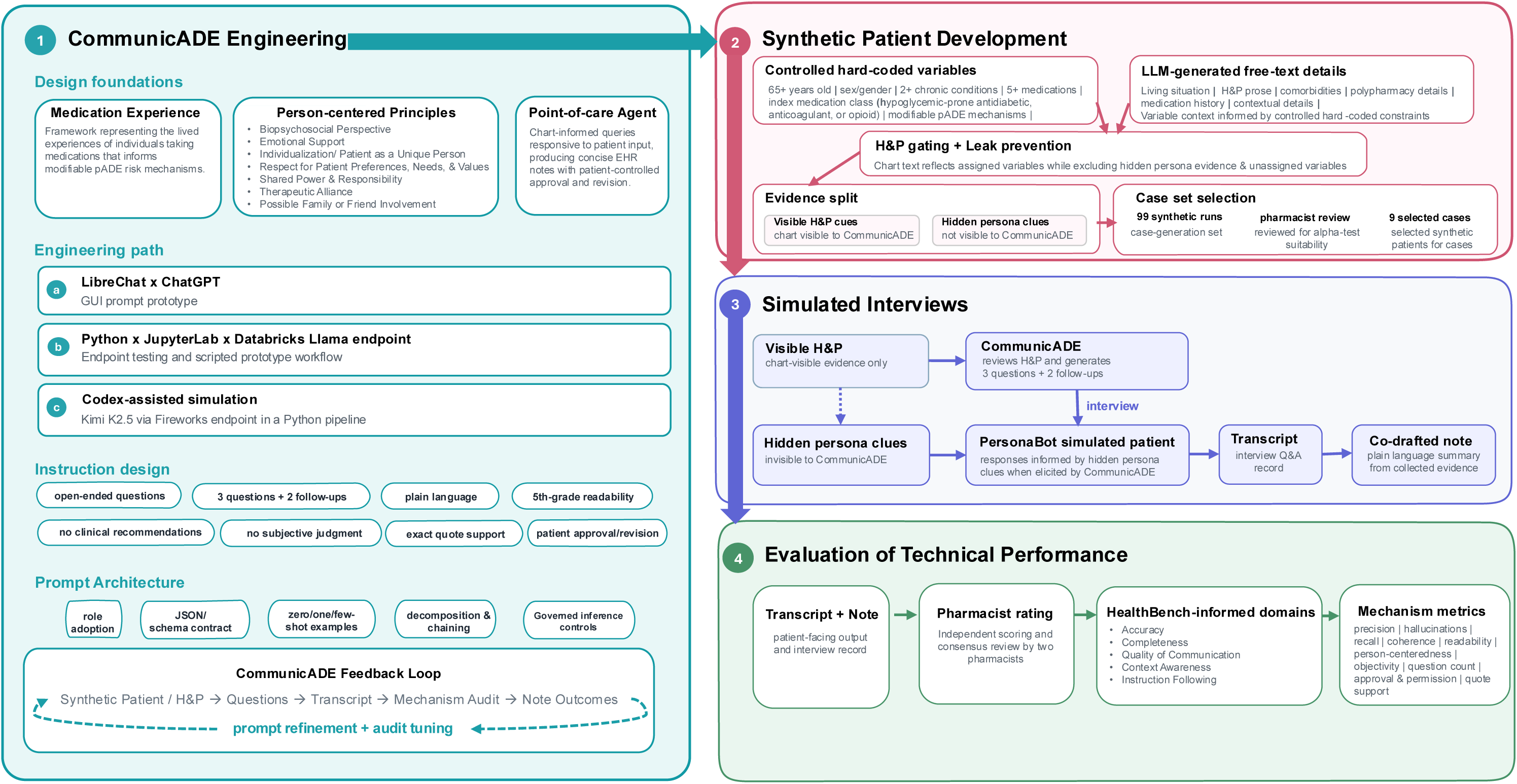
Description of CommunicADE Engineering, Synthetic Patient Development, Chatbot Simulation, and Evaluation Process. **ALT TEXT:** A graphical flow chart with descriptive text showing the sequential processes used to engineer CommunicADE, develop synthetic patients, conduct simulated interviews, and evaluate CommunicADE’s technical performance.

### CommunicADE Development

Developer prompts for CommunicADE were produced to: (1) help LLMs understand the goals of an interview with synthetic patients, especially the collection of information relevant to mediating mechanisms for modifying pADE risk from a synthetic History and Physical (H&P) and interviews; (2) develop a list of corresponding questions consistent with the interview goal; and (3) synthesize notes identifying modifiable determinant mechanisms with supporting patient-quotes.

The LibreChat^39^ browser graphical user interface (GUI) was used with ChatGPT models to explore concept development at Essentia Institute of Rural Health. The CommunicADE prototype was formally developed and refined at Geisinger using JupyterLab^40^ and Python 3.13^41^ to call internally governed, Databricks^42^-hosted, API endpoints for Llama 3.1 70B and 405B.^43^ The Python pipeline was ported externally with an OpenAI-compatible codex to Kimi K2.5 to improve performance and avoid barriers to reliably completing simulations (e.g., endpoint token-per-minute limits, and content guardrails for terms like “fentanyl” or “needles).”^44^

Engineering of the CommunicADE prototype was informed by the Medication Experience (MedXp) framework for identifying mediating mechanisms for modifying pADE risks (**Table 1**)^45–52^ as well as patient-centeredness models for patient communication. Additional prompts limited CommunicADE to asking five questions, using a person-centered approach, writing at a 5^th^ grade readability level, and refraining from clinical or health-related recommendations and subjective judgements.^53–54^ These constraints were added to improve CommunicADE’s performance on the five evaluation axes from OpenAI’s HealthBench.^34^ Full developer prompts are available in the supplementary material.

**Table 1.**
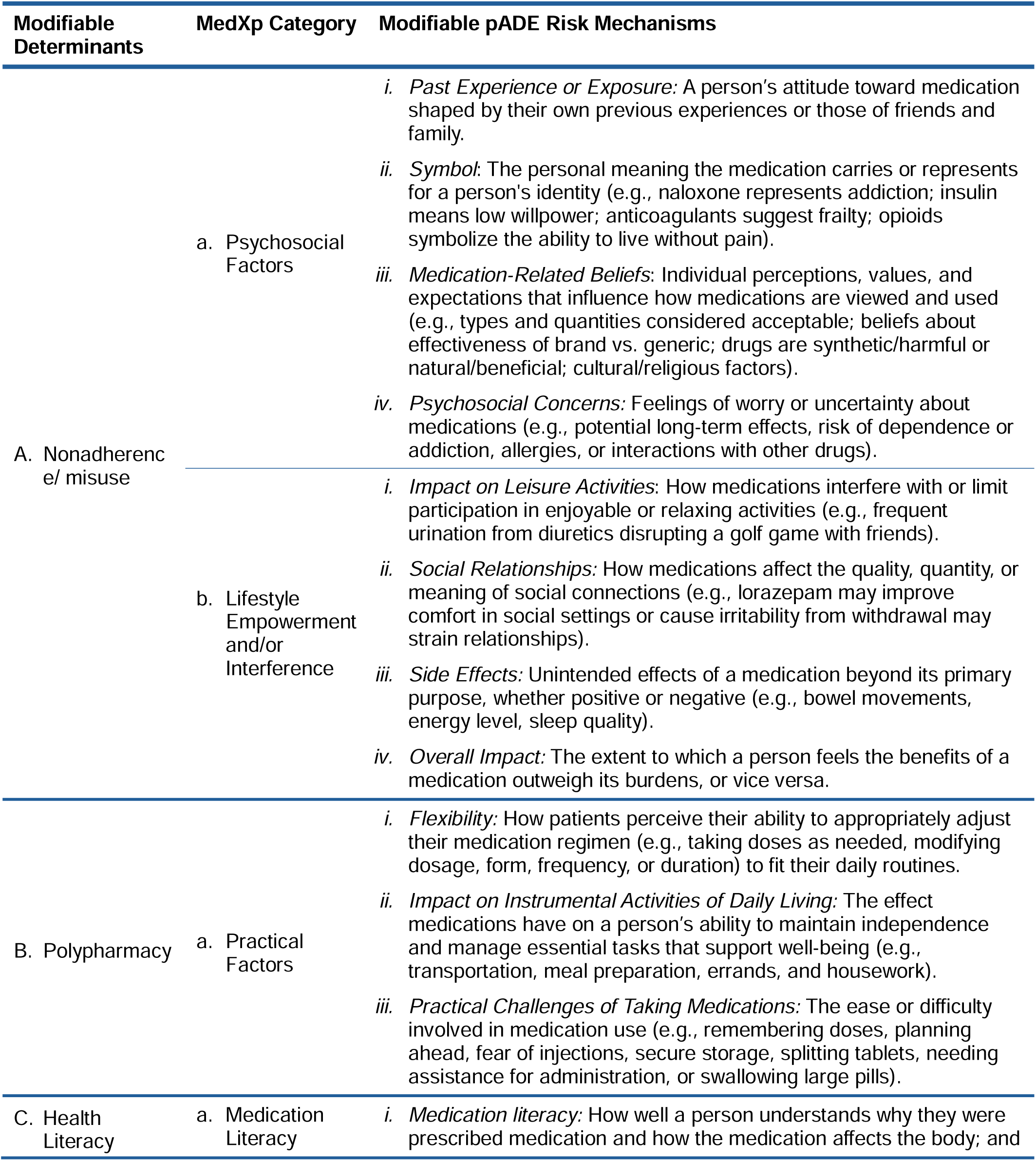
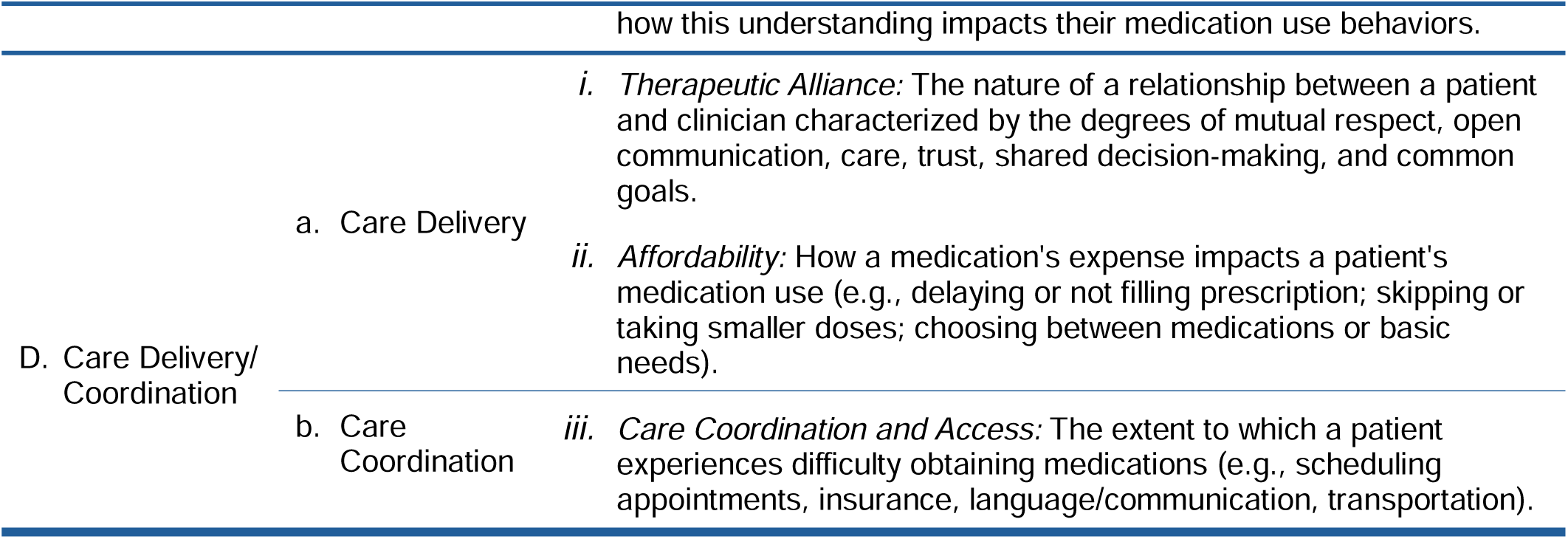
Modifiable preventable adverse drug event (pADE) risk mechanisms organized by the Medication Experience (MedXp) framework^12,20,45–52^.

Prompt engineering techniques were refined iteratively through trial and error. Purpose-specific prompts that were chained across discrete tasks using several techniques: hard-coded variables; zero-, one-, and few-shot examples; explicit persona framing (e.g.,“You are an expert medical scribe.”); and strict formatting (e.g., JSON schema). Gating techniques prevented mechanism leakage, unsupported inferences, clinical recommendations, and undesired content. To improve auditability and reduce hallucinations, prompts were grounded in source text from synthetic patients.

### Synthetic Patients

For the purposes of technical validation, our sampling strategy prioritized higher quality of synthetic patients used in the evaluation over having a higher quantity of synthetic patients. We employed a two-step approach. First, data for 99 synthetic patients (i.e., patients for whom CommunicADE was designed) were generated using Kimi K2.5. All synthetic patients were older adults (65+ years old) with a chronic comorbidity and polypharmacy (5+ prescriptions) hospitalized after presenting the emergency department with signs and symptoms of a suspected pADE. Three groups of 33 patients were assigned different high-risk ADE medication classes: (1) opioids; (2) anticoagulants; and/or (3) insulins or sulfonylureas.

In step two, a multi-rater assessment was conducted independently by 2 practicing pharmacists (AWO and DD) who selected 9 high-fidelity synthetic patients from the pool of 99 synthetic patients.^36,55^ Selection was based on consensus judgement of pharmacists on synthetic patient realism and appropriateness (e.g., eligibility criteria, fidelity to prompting). After selection, the pharmacists independently reviewed and detected 35 pADE mechanisms present in the 9 synthetic patients.

Each synthetic patient was assigned an H&P and a persona by Kimi K2.5; the former was accessible to CommunicADE and the latter was not. All synthetic patients were assigned modifiable pADE risk mechanisms tied to the high-risk ADE medications split between the H&P and persona. Patients could not have diagnoses of metastatic cancer, severe psychiatric disorders (i.e., bipolar disorder, schizophrenia, dementia) or present with an intentional opioid overdose. The H&P contained randomly generated information about the patient’s specific name, sex, age, comorbidities, and living situation (e.g., living alone in single-level home, senior community with elevator, dog at home). The H&P also included patient signs and symptoms consistent with their assigned high-risk prescription (e.g., bleeding with anticoagulant; hypoglycemia symptoms with insulin or sulfonylurea; unintentional respiratory depression, constipation, nausea/vomiting, withdrawal with opioid). Finally, 2+ mechanisms from MedXp categories (**Table 1**) related to medication-class specific issues commonly found in H&P (i.e., Side Effects, Affordability, Access, Instrumental Activities of Daily Living, Practical Challenges, Medication Literacy, Psychosocial Concerns, Flexibility, Impact on Leisure Activities, and Overall Impact) were assigned.

Synthetic patients’ personas and H&Ps shared overlapping patient characteristics (i.e., name, sex, age, comorbidities, and living situation), but distinct pADE risk mechanisms. Personas contained the pADE risk mechanisms often missing from the EHR despite being important opportunities to mitigate pADE risk (e.g., Past Experience/Exposure, Symbol, Medication-related Beliefs, Social Relationships, Practical Challenges of Taking Medications, Medication Literacy, Therapeutic Alliance, Care Coordination and Access). Persona profiles were not accessible to CommunicADE; a specific Kimi K2.5 agent served to validate prevention of data leakage. Thus, CommunicADE only made inferences from information collected in the simulated interviews. Personas were assigned 2+ mechanisms from different MedXp categories to ensure a desirable balance of mechanisms.

### Simulated Interviews

Refined prompts were programmed in Python^41^ to conduct and transcribe CommunicADE’s interview of the 9 high-fidelity synthetic patients as well as draft a corresponding note for the EHR. These data, which are available in the supplemental materials, were used to assess CommunicADE’s effectiveness and appropriateness.

CommunicADE initiated each simulated interview after reviewing the H&P to appropriately tailor person-centered questions for identifying pADE risk mechanisms. Interviews always began with an open-ended asking patients to tell the story of their hospitalization and how their medications may have played a part. The question intended to focus patients on medication-related details of their story rather than other factors less likely to be relevant in the intervention’s target population. CommunicADE’s next two questions were formulated to identify pADE risk mechanisms based on the contextual information from the H&P, and the final two questions were followed up on learnings from the preceding three questions to identify pADE risk mechanisms hidden in the persona and not evident from the H&P. CommunicADE then used the totality of the information collected to that point to generate a patient-friendly note (e.g., plain language, coherent structure, biopsychosocial approach) containing clinically meaningful information. The CommunicADE synthesized notes are available in the supplementary material.

### Variables

Each simulated interview was assessed along ten variables informed by OpenAI’s HealthBench five evaluation axes: accuracy, completeness, quality of communication, context awareness, and instruction following. **Table 2** describes each axis and operationally defines the study variables underlying the axes. HealthBench is an open-source evaluation framework for evaluating the usefulness, reliability, safety, and general performance of AI tools in healthcare contexts. More than 250 physicians from 60 countries representing 49 languages and 26 practice subspecialties assisted in the comprehensive benchmark’s development to optimize clinical utility.^34^

**Table 2.**
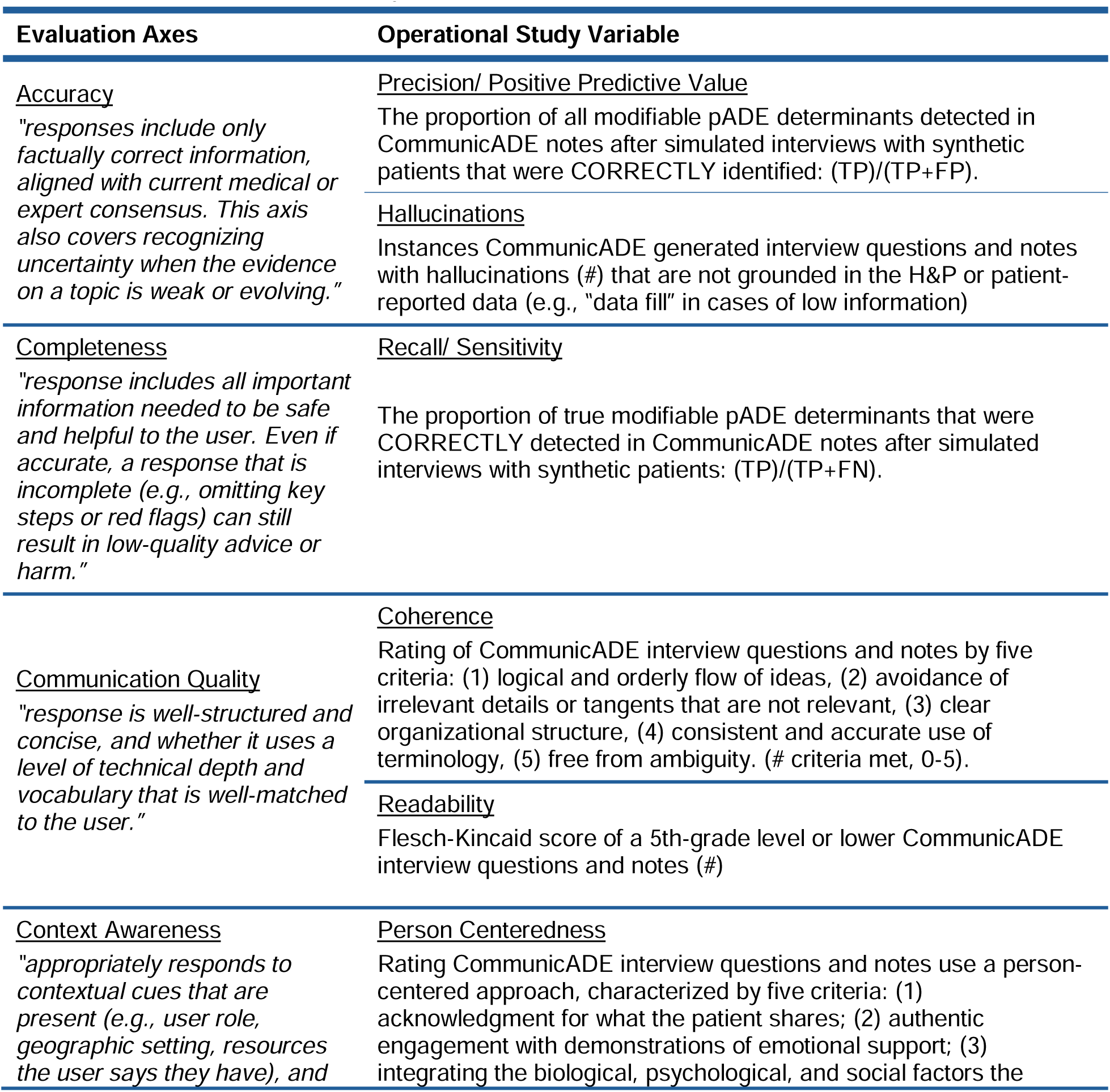
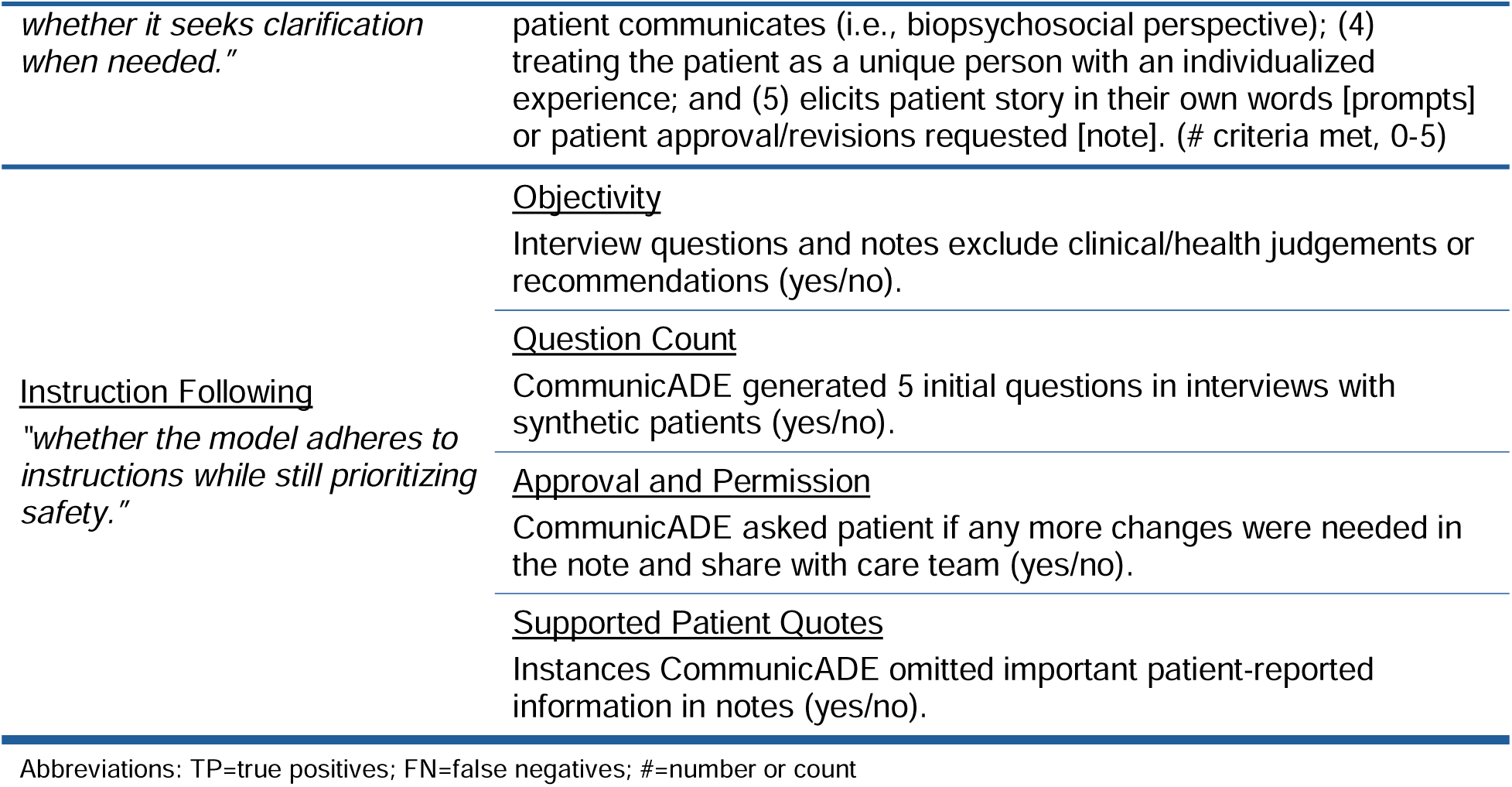
Operational Variables by OpenAI HealthBench Evaluation Axes^34^.

| Evaluation Axes | Operational Study Variable |
| --- | --- |
| <u>Accuracy</u><br>*“responses include only factually correct information, aligned with current medical or expert consensus. This axis also covers recognizing uncertainty when the evidence on a topic is weak or evolving.”* | <u>Precision/ Positive Predictive Value</u><br>The proportion of all modifiable pADE determinants detected in CommunicADE notes after simulated interviews with synthetic patients that were CORRECTLY identified: (TP)/(TP+FP). |
|  | <u>Hallucinations</u><br>Instances CommunicADE generated interview questions and notes with hallucinations (#) that are not grounded in the H&P or patient-reported data (e.g., “data fill” in cases of low information) |
| <u>Completeness</u><br>*“response includes all important information needed to be safe and helpful to the user. Even if accurate, a response that is incomplete (e.g., omitting key steps or red flags) can still result in low-quality advice or harm.”* | <u>Recall/ Sensitivity</u><br>The proportion of true modifiable pADE determinants that were CORRECTLY detected in CommunicADE notes after simulated interviews with synthetic patients: (TP)/(TP+FN). |
| <u>Communication Quality</u><br>*“response is well-structured and concise, and whether it uses a level of technical depth and vocabulary that is well-matched to the user.”* | <u>Coherence</u><br>Rating of CommunicADE interview questions and notes by five criteria: (1) logical and orderly flow of ideas, (2) avoidance of irrelevant details or tangents that are not relevant, (3) clear organizational structure, (4) consistent and accurate use of terminology, (5) free from ambiguity. (# criteria met, 0-5). |
|  | <u>Readability</u><br>Flesch-Kincaid score of a 5th-grade level or lower CommunicADE interview questions and notes (#) |
| <u>Context Awareness</u><br>*“appropriately responds to contextual cues that are present (e.g., user role, geographic setting, resources the user says they have), and* | <u>Person Centeredness</u><br>Rating CommunicADE interview questions and notes use a person-centered approach, characterized by five criteria: (1) acknowledgment for what the patient shares; (2) authentic engagement with demonstrations of emotional support; (3) integrating the biological, psychological, and social factors the |
| *whether it seeks clarification when needed."* | patient communicates (i.e., biopsychosocial perspective); (4) treating the patient as a unique person with an individualized experience; and (5) elicits patient story in their own words [prompts] or patient approval/revisions requested [note]. (# criteria met, 0-5) |
| *Instruction Following*<br>*"whether the model adheres to instructions while still prioritizing safety."* | <u>Objectivity</u><br>Interview questions and notes exclude clinical/health judgements or recommendations (yes/no). |
|  | <u>Question Count</u><br>CommunicADE generated 5 initial questions in interviews with synthetic patients (yes/no). |
|  | <u>Approval and Permission</u><br>CommunicADE asked patient if any more changes were needed in the note and share with care team (yes/no). |
|  | <u>Supported Patient Quotes</u><br>Instances CommunicADE omitted important patient-reported information in notes (yes/no). |

Abbreviations: TP=true positives; FN=false negatives; #=number or count

### Analysis

Two pharmacists (AWO and DD) independently produced or calculated measurements for all variables and met to resolve discrepancies. The attempt to reach consensus increases data trustworthiness (e.g., peer examination, confirmability audit), while preserving remaining disagreements to be reported.^56^ The resulting data for the 9 synthetic patients were summarized using descriptive statistics and compared across pharmacist evaluators. Results for variables were reported at both the “item-level” (i.e., individual pADE determinants) and the “patient-level.” Analyses were conducted separately for interview questions and note drafts. Inferential statistics were not used given data represented a census rather than a sample.

## RESULTS

### Synthetic Patient Characteristics

Among the 9 synthetic patients, 35 pADE risk mechanisms were detected by study pharmacists. The average number of pADE risk mechanisms per patient was 3.89 (sd=1.37). **Table 3** reports the CommunicADE’s performance across the five HealthBench criteria are described below.

**Table 3.** CommunicADE performance by HealthBench-informed variables^34^.

| HealthBench<br>Axes | Variables<br>(measure or unit) | Anticoagulant<br>s | Opioids | Antidiabetic<br>s | All |
| --- | --- | --- | --- | --- | --- |
|  |  |  | average (sd) |  |  |
| Accuracy | Precision ($TP/(TP+FP)$) | | | | |
|  | Patient-level (n=9) | 0.67 (0.58) | 0.92 (0.14) | 1 (0) | 0.86<br>(0.33) |
|  | Mechanism-level<br>(n=24) | 0.88 (0.33) | 0.91 (0.29) | 1 (0) | 0.92<br>(0.28) |
|  | Hallucinations (#) | 0 (0) | 0 (0) | 0 (0) | 0 (0) |
| Completeness | Recall ($TP/(TP+FN)$) | | | | |
|  | Patient-level (n=9) | 0.47 (0.42) | 0.43 (0.40) | 0.64 (0.38) | 0.51<br>(0.36) |
|  | Mechanism-level<br>(n=35) | 0.58 (0.49) | 0.71 (0.45) | 0.56 (0.50) | 0.63<br>(0.48) |
| Communication<br>Quality | Coherence (5-point scale) | 4 (0.63) | 4.5 (0.55) | 4.33 (0.82) | 4.28<br>(0.67) |
|  | Readability<br>(Flesch-Kincaid grade<br>level) | 4.3 (1.05) | 4.7 (0.55) | 4.65 (1.74) | 4.54<br>(1.16) |
| Context<br>Awareness | Person-Centeredness<br>(5-point scale) | 4.3 (0.82) | 4.5 (0.55) | 4.5 (0.55) | 4.44<br>(0.62) |
| Instruction<br>Following | Objectivity (0=no, 1=yes) | 0.67 (0.52) | 1 (0) | 0.67 (0.52) | 0.78<br>(0.43) |
|  | ≤5 Questions (0=no,<br>1=yes) | 1 (0) | 1 (0) | 1 (0) | 1 (0) |
|  | Approval & Permission<br>(0=no, 1=yes) | 1 (0) | 1 (0) | 1 (0) | 1 (0) |
|  | Quote Supports<br>Mechanism (0=no, 1=yes) |  |  |  |  |
|  | Patient-level (n=9) | 0.33 (0.58) | 1 (0) | 1 (0) | 0.78<br>(0.44) |
|  | Mechanism-level<br>(n=24) | 0.75 (0.43) | 1 (0) | 1 (0) | 0.92<br>(0.28) |

Note: Scoring of interviews and notes were assessed separately, but this table reports combined results of interviews and notes for the variables of coherence, readability, person-centeredness, and objectivity.
Interrater Rater Reliability: Two pharmacists achieved consensus on 115 of 117 independent judgements. The 2 unresolved disagreements produced the same quantitative scores but for different reasons in the variables of recall and coherence.
Abbreviations: sd=standard deviation; TP=true positive, FP=false positive, FN=false negative

### Accuracy

Of the 24 pADE risk mechanisms identified by CommunicADE, 92% were true positives (22 TPs across 8 patients). Two false positives (FPs) were identified by CommunicADE in different cohorts (1 anticoagulant, 1 antidiabetic), producing an average precision of 0.86 (sd=0.33) at the patient-level and 0.92 (sd=0.28) at the mechanism-level. Patient-level averages by cohort were 0.67 (sd=0.58) for anticoagulants, 0.92 (sd=0.14) for opioids, and 1 (sd=0) for antidiabetics, respectively. Mechanism-level averages by cohort were 0.88 for anticoagulants (sd=0.33), 0.91 (sd=0.29) for opioids, and 1 (sd=0) for antidiabetics, respectively. Hallucinations were absent from all interviews and notes.

### Completeness

Of 35 true pADE risk mechanisms, 37% were false negatives (13 FNs across 7 patients) for CommunicADE. CommunicADE’s average recall at the patient-level was 0.51 (sd=0.36) and 0.63 (sd=0.48) at the mechanism-level. The patient-level averages by cohort was 0.47 (sd=0.42) for anticoagulants, 0.43 (sd=0.40) for opioids, and 0.64 (sd=0.38) for antidiabetics, respectively. The mechanism-level average by cohort was 0.58 for anticoagulants (sd=0.49), 0.71 (sd=0.45) for opioids, and 0.56 (sd=0.50) for antidiabetics, respectively.

### Communication Quality

CommunicADE’s average coherence on a 5-point scale was 3.78 for interview questions. Cohort averages for interview questions was 3.67 for anticoagulants, 4 for opioids, and 3.67 for antidiabetics, respectively. Coherence downgrades were for non-orderly flow of logic/ideas between questions (7 of the 9 interviews) and organizational structure from odd wording (3 of 9 interviews). The average coherence for note drafts was 4.78, with cohort averages of 4.33 for anticoagulants, 5 for opioids, and 5 for antidiabetics. The average coherence combining interview questions and note drafts was 4.28 (sd=0.67).

The average Flesch-Kincaid reading grade level for CommunicADE interview questions was 5.34. Cohort averages for interview questions was 4.83 for anticoagulants, 5.03 for opioids, and 6.17 for antidiabetics, respectively. The average reading grade level for note drafts was 3.73. Cohort averages were 3.73 for anticoagulants, 4.33 for opioids, and 3.13 for antidiabetics. The average reading grade level combining interview questions and note drafts was 4.54 (sd=1.16).

### Context Awareness

The average person-centeredness rating for CommunicADE on a 5-point scale was 3.89 for interview questions. Cohort averages for interview questions was 3.67 for anticoagulants, 4 for opioids, and 4 for antidiabetics, respectively. All interviews were absent of questions demonstrating evidence of emotional support for the patient, and 1 interview asked a question that missed or failed to acknowledge information provided by the patient in a preceding response. The average rating for note drafts in all cohorts was 5. The average person-centeredness rating combining interview questions and note drafts was 4.44 (sd=0.62).

### Instruction Following

All of CommunicADE’s interview questions were free of non-objective statements, but the same was true for only a little over half (56%) of draft notes. Opioid cohort draft notes did not contain non-objective statements, but the anticoagulant and antidiabetics cohorts each had 2 notes with non-objective or clinical statements (e.g., “your blood was too thin because you changed your warfarin dose on your own”). Thus, the combined percentage of interview questions and draft notes without objective statements was 78%.

All patients were asked five questions in interviews and all draft notes asked patients for their approval and permission to share with the healthcare team in the EHR. All CommunicADE notes contained patient quotes, of which 92% were appropriately grounded in the pADE risk mechanism detected. Two anticoagulants notes contained quotes misaligned with the reported pADE risk mechanism.

## DISCUSSION

To our knowledge, there are no LLM chatbots engineered to collect and facilitate use of patients’ medication-related values, preferences, beliefs, and goals to facilitate reduction of pADE risk. In this evaluation using synthetic patients informed by benchmarks from OpenAI’s HealthBench framework, CommunicADE performed well across multiple axes. CommunicADE’s technical performance indicates readiness for a feasibility pilot with real-world clinical encounters. Findings suggest that the prototype can add utility through precise identification of modifiable medication risks absent from H&Ps. CommunicADE displayed levels of communication quality (i.e., coherence and readability) and context awareness (i.e., person-centeredness) suggesting neutral to positive patient experience impact as well as limited risk of hallucinations and inappropriate clinical judgement. Sizable opportunities remain for improving CommunicADE’s identification of all pADE risk mechanisms patients possess (i.e., completeness/recall).

### Accuracy

High accuracy in identifying modifiable pADE mechanisms is critical for CommunicADE to be effective and sustainably utilized. Patients are likely to have low tolerance for questions or initial draft notes incongruent with their experiences, even if patients can iterate revisions of notes. Similarly, clinicians need to be confident that the pADE mechanisms identified are not false positives that could waste their time, effort, resources, or even cause harm. CommunicADE’s positive predictive value (0.916) and absence of hallucinations suggest an aspect of readiness for piloting the prototype with real patients. Sample size limited ability to detect statistically significant performance differences between medication class cohorts; this will be closely monitored when CommunicADE is pilot-tested with real patients.

### Completeness

The CommunicADE prototype performance in recall was less favorable than its precision. While higher recall is always preferable, the application increases the likelihood of identifying modifiable pADE mechanisms often missed or absent from the EHR. Furthermore, the patient’s ability to revise draft notes generated by CommunicADE offer a way to improve completeness that was not assessed in our evaluation. Reasons for why recall was poorer among the anticoagulant and opioid cohorts are unclear and close monitoring to mitigate for care quality differences across high-risk medications.

### Communication Quality

CommunicADE scored well on communication quality, exhibiting good coherence representative of the information provided by patients. The average coherence score of interview questions (x□=3.78) was worse than note drafts (x□=4.78), which may be attributable to differences in their functional purpose and length limits. The most frequently coherence downgrades of questions due to disordered flow of ideas/logic may be associated by the exploratory purpose of the interviews combined with the limitations of five questions leading to non-linear jumps in topics. Similarly, the coherence downgrades from odd wording of questions may reflect CommunicADE’s attempt to probe for multiple underlying constructs within question limits. The coherence downgrades for note drafts were for inaccurate use of terminology (2/9 notes) and inclusion of irrelevant details (1/9 notes). Both issues may be traced to CommunicADE’s difficulty discriminating between some similar pADE risk mechanisms (e.g., medication negatively impacting a *Social Relationship* versus being a potential *Symbol* of why they have poor relationships), suggesting the need for more refinement of pADE risk mechanism operational definitions or additional few-shot prompting (e.g., additional training with examples that more clearly highlight distinctions).

The average Flesch-Kincaid Grade Level readability score of interview questions (5.34) and note drafts (3.73) were below the recommended reading level for older adults.^57^ Readability is a key component of health literacy, especially among older adults,^58^ that can prevent patients from obtaining, understanding, and making informed medication use choices and lead to pADEs.^59–63^ The result may reflect the different purposes and length constraints put on interview questions, which would require longer sentences (e.g., conditional phrasing, embedded clauses) and bigger words (e.g., multi-syllabic words) to explore and elicit more useful responses. These characteristics would lead to higher Flesch-Kincaid grade levels, which are mostly a derivative of sentence length and syllables per word averages.

### Context Awareness

CommunicADE met most person-centeredness criteria in interview questions (x□=3.89), and all criteria for draft notes (x□=5). The lack of emotional support apparent in all interview questions may again be attributable to exploratory purpose question number limits. A way to address this gap may be for CommunicADE to express emotional support for the patient with empathetic language prior to asking the first question. CommunicADE can also be engineered to supportively acknowledge patient responses before preceding to the next question.

The full scores achieved by CommunicADE in all note drafts suggest successful incorporation of person-centered principles engineered into the application. The findings are also consistent with evidence that chatbots have rated favorably for empathy in patient interactions and often outperform clinicians completing the same task.^24, 64–65^

### Instruction Following

All of CommunicADE’s interviews contained only five questions, which were free of non-objective statements. Of 9 draft notes, 5 possessed 1+ statement of subjectivity. Additional prompt engineering is necessary to eliminate non-objective statements in draft notes to ensure CommunicADE functions solely as a supplemental tool that augments patient-clinician interactions with low risk of potential harm.

All patients were asked five questions in interviews and all draft notes ended with a statement asking patients for their approval and permission to share with the healthcare team in the EHR. This is a key component of CommunicADE function and value, because it gives patients the opportunity to align what is written in draft with their intended meaning and directly approve what is communicated to the care team. Thus, CommunicADE draft notes may help reduce the estimated 21%-90% of EHR notes containing documentation errors of patient-reports,^66–69^ which introduce clinical, safety, trust erosion, and legal risks.^70–71^

While all CommunicADE draft notes provided patient-reported quotes, a small proportion didn’t sufficiently support the corresponding mechanism. While patients would have the opportunity to correct, substitute, or improve the supporting quote, higher performance in this area may improve the confidence, utility, and effectiveness of the application in the eyes of both patients and clinicians.

### Limitations and Future Research

Several limitations of this technical validation study should be considered alongside our findings. Our evaluation of CommunicADE utilized synthetic patients, which is an emerging methodology that can produce high-fidelity simulations between patients and clinicians,^36^ but may omit key factors or introduce non-reflective biases (e.g., overly optimistic or simplified) that are different from real-world assessment settings.^22^ For these reasons, we employed the two-step approach involving a multi-rater assessment by practicing clinicians to verify for eligibility criteria, realism, and appropriateness. The tradeoff was a smaller sample size preventing detection of statistically significant differences between medication cohorts or other variables (e.g., health literacy levels, demographics), but these objectives are better evaluated in studies involving actual patients.^33^ A real world pilot will also be able to better evaluate CommunicADE’s performance with a patient who doesn’t follow instructions, provides answers unrelated to the question asked, or may respond in a hostile manner. Finally, our study is representative of CommunicADE’s performance with a single open-sourced LLM at one point in time. Given the rapid advances in artificial intelligence, it’s possible that CommunicADE’s performance may differ when using other LLMs.^72^ While we anticipate the use of subsequent LLM versions will improve rather than degrade CommunicADE’s overall performance, continual monitoring across LLM versions and types will be important.

## CONCLUSION

This paper introduces a novel LLM application named CommunicADE designed to support clinicians in the secondary prevention of pADEs for the patient population at highest risk for these events: older adults (≥65 years) with comorbidities and polypharmacy that includes treatment with opioids, anticoagulants, and/or hypoglycemic-prone antidiabetic agents (i.e., insulins, sulfonylureas). More specifically, CommunicADE aims to improve the timely collection, organization, and transmission of important contextualized patient-reported medication use information to the clinician, helping patients to feel heard, safe, and cared for, and enabling shared decision making.

We describe the sequential processes used to engineer CommunicADE as well as our approach to validating the application’s technical performance by conducting simulated interviews with high-fidelity synthetic patients representative of its target patient population. CommunicADE’s strongest performance was in accuracy (precision, hallucinations), communication quality (coherence, readability), and context awareness (person-centeredness). Completeness (recall) and instruction following (objectivity, pADE mechanism/quote alignment) show room for improvement. CommunicADE’s technical performance in this study indicates readiness for a feasibility pilot with real-world clinical encounters.

## Data Availability

All data produced in the present study are available upon reasonable request to the authors

## ACKNOWLEDGEMENTS

The authors thank Jeff Ferber and Michelle Sikkink (Essentia Health) for their technical expertise, general assistance, and/or guidance over the course of the study. OpenEvidence was consulted to supplement our review of the literature and ensure comprehensive contextualization of our findings. Microsoft CoPilot was used to check for errors of formatting in the references. OpenAI’s Sora and Microsoft CoPilot were utilized to generate imagery used in Figures 1 and 2, respectively.

## CREDIT STATEMENT

Dobbins: Conceptualization, Methodology, Software, Validation, Formal Analysis, Investigation, Resources, Writing – Original Draft, Writing – Review & Editing, Visualization. Russell: Conceptualization, Methodology, Writing – Review & Editing. Gunther: Writing - Review & Editing, Visualization. Shetty: Conceptualization, Writing – Review & Editing. Shomali: Conceptualization, Writing – Review & Editing. Vawdrey: Conceptualization, Writing – Review & Editing; Waring: Conceptualization, Resources, Writing – Review & Editing. Whary: Writing – Review & Editing, Project Administration, Visualization. Wong: Conceptualization, Methodology, Writing – Review & Editing; Wright: Conceptualization, Writing – Review & Editing. Olson: Conceptualization, Methodology, Formal Analysis, Investigation, Resources, Writing – Original Draft, Writing – Review & Editing, Visualization, Supervision, Funding Acquisition.

